# Free-Cog reformulated: analyses as independent or stepwise tests of cognitive and executive function

**DOI:** 10.1101/2023.01.29.23285153

**Authors:** Andrew J Larner, Alistair Burns

## Abstract

Free-Cog is a recently described, hybrid screening instrument incorporating tests of cognitive and executive function. In this study, Free-Cog was reformulated as separate tests of cognitive function and executive function to examine whether this might improve screening accuracy for cognitive impairment (dementia and mild cognitive impairment) compared to the standard, unitary Free-Cog. The two separate tests, designated “Free-Cog-Cog” and “Free-Cog-Exec,” were either combined using the Boolean logical “AND” and “OR” operators (serial and parallel combination) or used to construct a stepwise decision tree. Serial combination improved specificity and positive predictive value whereas parallel combination improved sensitivity, findings typically observed when applying these operators. Stepwise application identified groups with high and low probability of cognitive impairment but failed to adequately differentiate those in the intermediate uncertain diagnosis group. Although the dataset used was relatively small, the findings of this study suggest little benefit for reformulations of Free-Cog compared to the standard, unitary instrument.

## 1. Introduction

Identification of patients with cognitive impairment often involves the administration of cognitive screening instruments, many of which are available.^1,2^ Each screener has its particular advantages and limitations, and hence clinician choice and preference may vary. As the dementia syndrome by canonical definition includes functional limitation as well as cognitive impairment,^3^ tests examining function may be included in such screening assessments, and again many such instruments are available. Combining the outcomes of more than one test may add to the utility of screening procedures for cognitive impairment. This may involve not only more than one cognitive test but also a combination of cognitive and other tests, including functional assessment.^4,5^

Another approach to cognitive screening is stepwise case finding in which only those participants screening positive or negative on an initial test are then subjected to further testing. Examples of this approach include the UK Dementia CQUIN (Commissioning for Quality and Innovation) single question screener as a prelude to further referral or testing,^6^ and a similar single question designated the progressive forgetfulness question (PFQ).^7^ Staged, sequential testing strategies may be structured as decision trees incorporating successive dichotomous actions. An example of this approach in the field of cognitive screening instruments is the cognitive disorders examination or Codex,^8^ a two-step decision tree for diagnostic prediction which incorporates two sequential tests: in step 1 the results of three-word recall and spatial orientation tests are trichotomised (both tests normal, both tests abnormal, only one test normal), with the intermediate group then subjected to step 2, a simplified clock drawing test with these results dichotomised (normal, abnormal) resulting in four terminal nodes with different probabilities of dementia diagnosis. Other, existing screening instruments may also be reformulated as decision trees, e.g. the Mini-Cog.^9^

Free-Cog is a recently introduced hybrid short cognitive screening instrument incorporating tests of both cognitive function and executive function (item content shown in Table 1; score range 0-30, higher scores better).^10,11^ Initial studies have suggested that Free-Cog is an acceptable and efficacious instrument for the identification of cognitive impairment,^10,12^ whose metrics compare favourably with other brief cognitive screening instruments.^10,12,13^ Free-Cog is available free of copyright in both standard and abridged forms suitable respectively for face-to-face^10^ and remote (telephone, video)^10,14^ testing.

**Table 1:** Item content of Free-Cog.

|  | **“Cognitive Function”** |
| --- | --- |
| General Knowledge | 1 |
| Orientation: Time | 3 |
| Orientation: Place | 3 |
| Registration | 0<br>(5 words) |
| Calculation | 3 |
| Attention/Concentration | 2 |
| Memory recall: | 5<br>(Recall of previously presented 5 words) |
| Verbal fluency in 1 minute | 1<br>(semantic: animals) |
| Language: Naming | 2 |
| Language: Repetition | 1 |
| Language: write a sentence | 1 |
| Visuospatial abilities: Wire (Necker) cube | - |
| Visuospatial abilities: Clock drawing test | 3 |
|  | **“Executive Function”** |
|  | 5 (questions relating to social function, travel, home, emergency, and care function) |
| **Total Score** | **30** |

The hybrid nature of Free-Cog permits its reformulation in ways which might enhance its value as a screening tool for cognitive impairment. The purpose of this study was to reformulate Free-Cog and to examine test outcomes using two distinct strategies, viz.:

### Strategy 1

Free-Cog item content (Table 1) was reformulated as two separate tests, designated “Free-Cog-Cog” (range 0-25, higher scores better) and “Free-Cog-Exec” (range 0-5, higher scores better). The outcomes of these two tests were then combined using the Boolean logical operators “AND” and “OR” (also known as series and parallel combination respectively). All study participants were thus classified according to the results of both tests.

### Strategy 2

Free-Cog was reformulated as a decision tree, as per the cognitive disorders examination Codex (Figure 1). Hence outcomes were initially trichotomised according to the results of Free-Cog-Cog into high probability normal (no cognitive impairment) and abnormal (cognitive impairment) groups with an intermediate uncertain group. The latter group was then further classified according to the results of Free-Cog-Exec scores. Hence, unlike Strategy 1, only a subset of all participants was classified by stepwise application of both tests.

**Figure 1:**
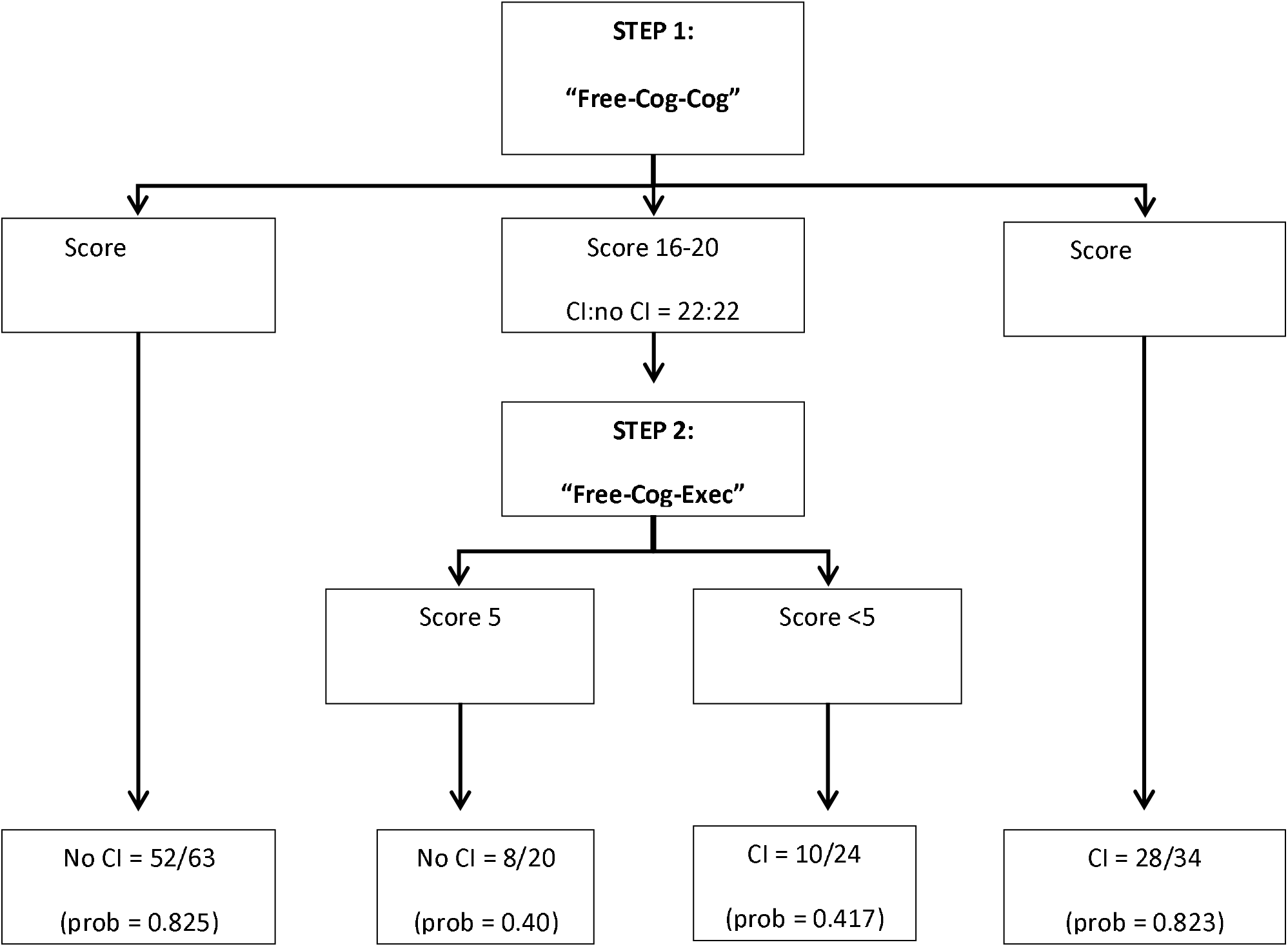
Free-Cog rewritten as a stepwise decision tree, incorporating results from reanalysis of the index Free-Cog study^12^ dataset.

## 2. Material and methods

The dataset from a pragmatic prospective screening test accuracy study examining Free-Cog (November 2017-October 2018 inclusive) in consecutive new patients referred to a neurology-led dedicated cognitive function clinic based in a regional neuroscience centre^12^ was re-interrogated. Subjects gave informed consent and the original study protocol was approved by the institute’s committee on human research (Walton Centre for Neurology and Neurosurgery Approval: N 310).

Patient assessment comprised semi-structured history enquiring about cognitive symptoms and functional performance, with collateral history from a reliable and knowledgeable informant where possible. All patients underwent neuroradiological examination (brain CT) with interval MR imaging in some cases. Formal neuropsychological assessment was pursued in some cases. Administration of Free-Cog occurred on the same day as, but separate from, the cross-sectional assessment. Criterion diagnosis of dementia, MCI, or subjective memory complaint (SMC), was by judgment of an experienced clinician based on diagnostic criteria (DSM-IV for dementia; Petersen for MCI) but did not use Free-Cog score in order to avoid review bias. STARDdem guidelines for reporting diagnostic test accuracy studies in dementia were observed.^15^

### 2.1 Strategy 1: combining tests

Initial Free-Cog analysis was as a single, global, unitary screening instrument (score range 0-30, higher better), using the cut-off previously defined in this patient cohort by maximal Youden index (≤22/30).^12^ From the 2×2 contingency table, standard test outcome measures were calculated: sensitivity (Sens), specificity (Spec), positive and negative predictive values (PPV, NPV), and overall test accuracy (Acc).

Free-Cog was then analysed as sequential screening instruments: “Free-Cog-Cog” (score range 0-25, higher better; Figure 2A), and “Free-Cog-Exec” (score range 0-5, higher better; Figure 2B). The optimal cut-off for each test was defined by maximal Youden index. These tests were then combined in series (“AND”) or in parallel (“OR”) to generate 2×2 contingency tables from which the same outcome measures as for the global Free-Cog test could be calculated and compared.

**Figure 2:**
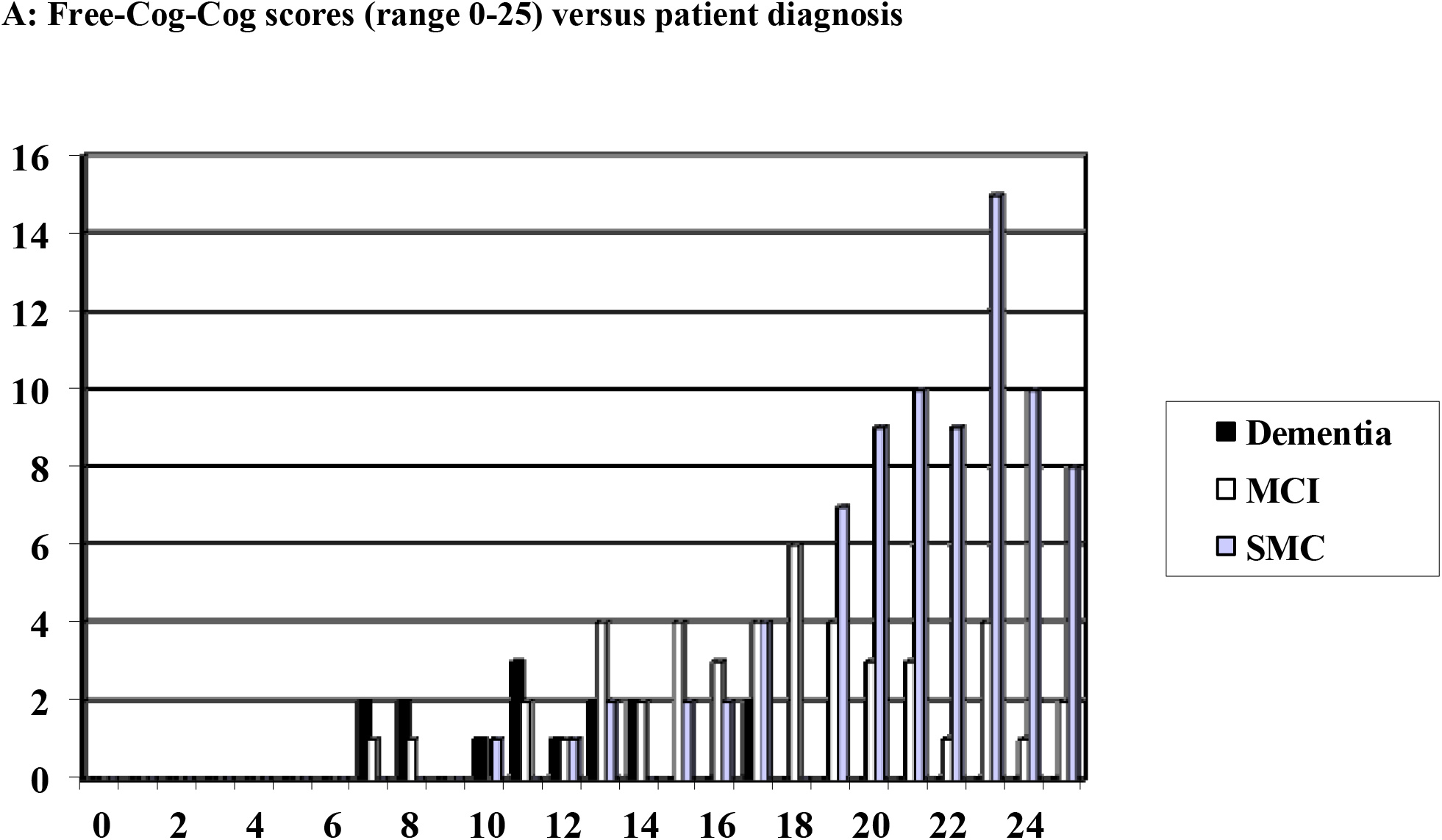
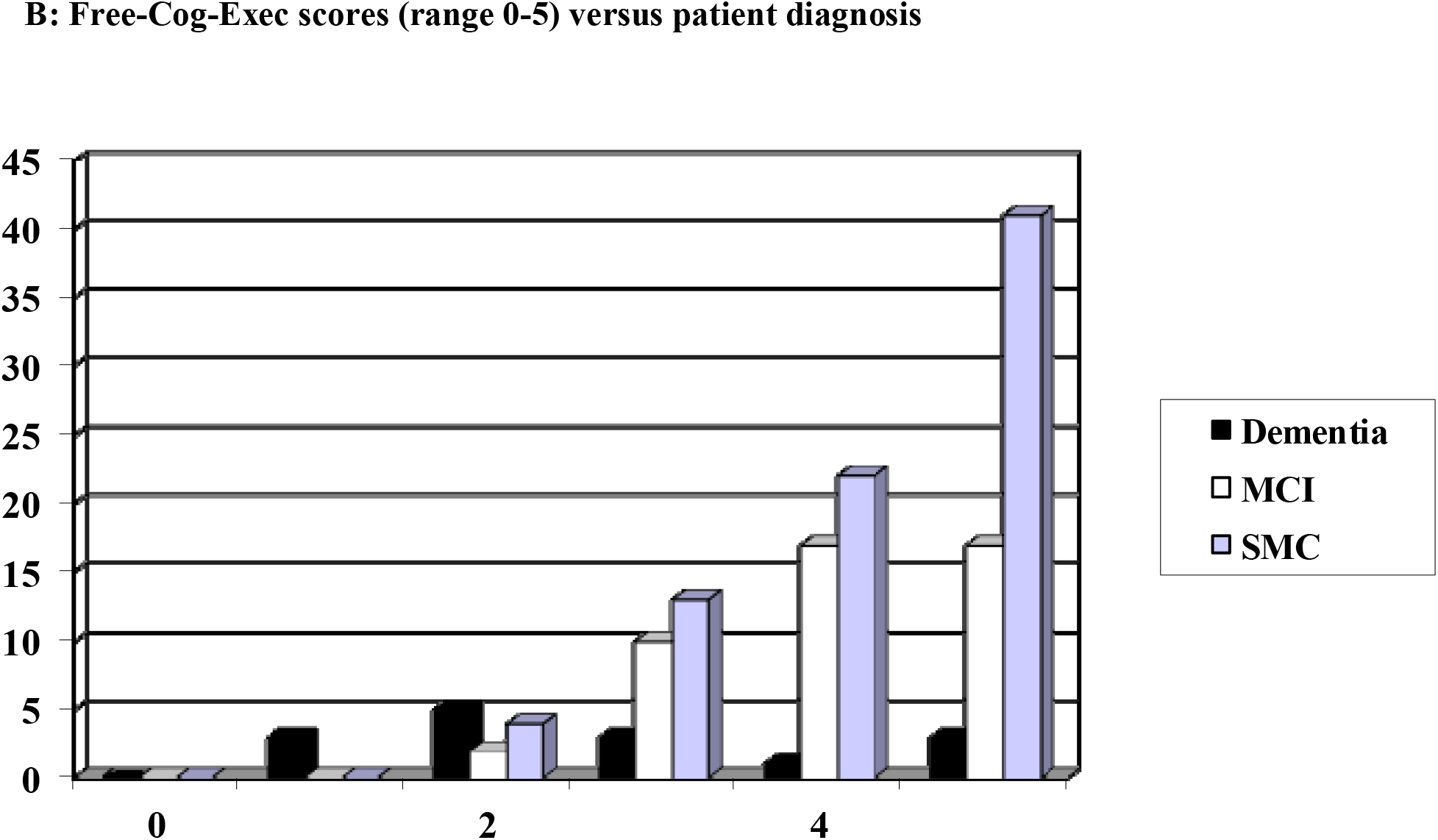

### 2.2 Strategy 2: stepwise decision tree

Results of Free-Cog-Cog were initially trichotomized (Figure 1). Results falling near a dichotomous cut-off (in this case ≤18/25) are those most likely to be uncertain or error-prone,^16^ whereas those far below or far above the cut-off are most likely to give a correct diagnosis, in this instance of cognitive impairment or no cognitive impairment respectively. Because of the rightward, negative skewed distribution of Free-Cog-Cog scores (Figure 2A), the trichotomisation step was not anticipated to produce groups of equal size.

The group of patients with Free-Cog-Cog scores falling close to the threshold, and hence with uncertain diagnosis, were then dichotomised using their Free-Cog-Exec scores to see if this improved classification accuracy.

## 3. Results

Of the 141 patients included in the study (F:M = 61:80, 43% female; age range 28-88 years, median age 62 years), criterion diagnoses were dementia (n = 15), MCI (n = 46), and SMC (n = 80).

### 3.1 Strategy 1: combining tests

The optimal cut-off for Free-Cog-Cog in this patient cohort was found to be ≤18/25 and for Free-Cog-Exec it was ≤4/5. Hence combining these tests in the “AND” condition required both Free-Cog-Cog ≤18/25 and Free-Cog-Exec ≤4/5 for the diagnosis of cognitive impairment (dementia or MCI). Combining these tests in the “OR” condition required either Free-Cog-Cog ≤18/25 or Free-Cog-Exec ≤4/5 for the diagnosis of cognitive impairment, hence a more liberal criterion than the “AND” condition.

Comparing the results of “AND” and “OR” combination of Free-Cog-Cog and Free-Cog-Exec with the results for the standard, unitary, global Free-Cog^12^ (Table 2), it was evident that the more stringent “AND” condition gave better outcomes for Spec and PPV than global Free-Cog, whereas the more liberal “OR” condition gave better Sens and matching NPV.

**Table 2:** Free-Cog for diagnosis of cognitive impairment (dementia and MCI) versus no cognitive impairment (SMC): comparison of standard summary measures of discrimination (with 95% confidence intervals) using standard Free-Cog versus serial and parallel combination of separate Free-Cog-Cog and Free-Cog-Exec tests.

| **Test strategy** | **Sens** | **Spec** | **PPV** | **NPV** | **Acc** |
| --- | --- | --- | --- | --- | --- |
| Standard unitary Free-Cog ($\leq 22/30$) | 0.68<br>(0.51-0.86) | 0.81<br>(0.73-0.90) | 0.73<br>(0.62-0.85) | 0.78<br>(0.69-0.87) | 0.76<br>(0.69-0.83) |
| Free-Cog-Cog ($\leq 18/25$)<br>“AND”<br>Free-Cog-Exec ($\leq 4/5$) | 0.58<br>(0.46-0.71) | 0.88<br>(0.80-0.95) | 0.78<br>(0.66-0.90) | 0.74<br>(0.65-0.83) | 0.75<br>(0.68-0.82) |
| Free-Cog-Cog ($\leq 18/25$)<br>“OR”<br>Free-Cog-Exec ($\leq 4/5$) | 0.82<br>(0.72-0.91) | 0.49<br>(0.38-0.60) | 0.54<br>(0.44-0.65) | 0.78<br>(0.67-0.90) | 0.63<br>(0.55-0.71) |

### 3.2 Strategy 2: stepwise decision tree

Trichotomisation according to Free-Cog-Cog scores of ≥21/25 and ≤15/25 identified groups with high probabilities of no cognitive impairment (0.825) and cognitive impairment (0.823) respectively (Figure 1, Step 1).

In the intermediate group, patients with Free-Cog-Cog scores in the range 16/25-20/25, criterion diagnoses were equally balanced between presence and absence of cognitive impairment. Looking at the Free-Cog-Exec scores in this group (Figure 1, Step 2) failed to enhance classification, because at neither of the possible Free-Cog-Exec cut-off points (<5/5 or <4/5; no patient in this intermediate group scored <3/5) did those with cognitive impairment perform worse. That is, there were more patients with cognitive impairment in the group with higher (i.e. better) Free-Cog-Exec scores, and more patients with no cognitive impairment in the group with lower (i.e. worse) Free-Cog-Exec scores.

## 4. Discussion

This study examined the effects on test outcomes of reformulations of the Free-Cog instrument, made possible by the novel hybrid nature of this test. Free-Cog-Cog and Free-Cog-Exec were either combined or applied as a stepwise decision tree.

In Strategy 1, combining Free-Cog-Cog and Free-Cog-Exec using Boolean logical operators, the outcomes were those generally observed with the use of these operators: “AND” has better Spec and PPV than the global test, avoiding false positives; whereas “OR” has better Sens and NPV, avoiding false negatives. Hence the choice of combination strategy depends on a clinician’s priorities, whether to avoid false positives (high specificity) or to avoid false negatives (high sensitivity).

In Strategy 2, using Free-Cog-Cog and Free-Cog-Exec as a stepwise decision tree, the initial trichotomisation according to Free-Cog-Cog scores (Step 1) identified groups with high and low probability of cognitive impairment and an intermediate uncertain group. However, the dichotomisation of this latter group according to Free-Cog-Exec scores (Step 2) did not improve classification and hence screening utility. The limited score range of Free-Cog-Exec (0-5) might contribute to this finding, likewise the relatively simple questions asked, such that scores are ceiling were found in many of those with a criterion diagnosis of cognitive impairment (20/61).

The study limitations were those endemic to any clinic-based study. The selected study population had a relatively high prevalence of dementia and MCI compared to patient cohorts in community-based (e.g. primary care, population) cohorts. The study cohort was quite small (<150), necessitating pooling of dementia and MCI as a summary cognitive impairment group, and resulting in broad 95% confidence intervals. Use of cross-sectional clinical diagnoses as reference standard although idiomatic of day-to-day practice is potentially liable to error without delayed verification (e.g. no neuropathological data were available). All these factors might limit the generalizability of the study findings.

Test reformulations may be more cumbrous and complex, and their use would therefore be justified only if classification accuracy outcomes were better. The evidence from this study, admittedly with a relatively small patient cohort and from a specialist setting, do not encourage the view that these manipulations offer any major benefit to the standard global Free-Cog, but this point might still be worthy of examination in larger patient cohorts from other clinical settings with different prevalence of cognitive impairment.

## Data Availability

All data produced in the present study are available upon reasonable request to the authors

